# Effects of high-intensity interval training in patients with coronary artery disease after percutaneous coronary intervention: a systematic review and meta-analysis

**DOI:** 10.1101/2020.06.02.20119958

**Authors:** Xinyue Zhang, Dongmei Xu, Guozhen Sun, Zhixin Jiang, Jinping Tian, Qijun Shan

## Abstract

**Background:** High-intensity interval training, for its characteristic of short-time high oxygen-consumption exercise interphase with periods of low-intensity training or rest for recovery, is easier to persist and execute in cardiac rehabilitation. However, it is little known whether HIIT program has an advantageous effect on patients after percutaneous coronary intervention or not.

**Methods:** Randomized controlled trials (RCTs) focusing on HIIT program in patients after PCI were searched in Cochrane Library, Web of Science Core Collection, EMbase, PubMed, China National Knowledge Infrastructure (CNKI) and SinoMed from the inception to March 24, 2020. Two reviewers conducted the literature retrieval, data extraction, and quality assessment independently. Standard Mean difference (SMD) and 95% confidence intervals (CI) were performed to summarize the effect sizes.

**Results:** 6 RCTs (247 patients) met the criteria. HIIT program had a statistically significant effect on raising left ventricular ejection function (LVEF) (SMD=0.38, 95%CI[0.03, 0.73], p=0.03), VO_2peak_ (SMD=0.94, 95%CI[0.61, 1.28], p<0.01), as well as improving the serum level of high-density lipoprotein (SMD=0.55, 95%CI[0.06, 1.03], p=0.03) and late luminal loss (SMD=−0.65, 95%CI[−1.07, −0.23], p<0.01). But HIIT had no prominent effect on improving heart rate (SMD=−0.04, 95%CI[-0.29, 0.21], p=0.73).

**Conclusions:** HIIT program might be favorable for CAD patients after PCI by improving cardiopulmonary function, such as LVEF and VO_2peak_, as well as reducing late luminal loss in per stented arteries. Nevertheless, HIIT had no advantage for adjusting heart rate. More researches with rigorous methods are warranted to explore the controversy about lipid profiles.

## 1. Introduction

Percutaneous coronary intervention (PCI) has become the most effective treatment of coronary artery disease (CAD). Although PCI plays a significant role in decreasing the rate of vascular restenosis and recurrent ischemia, the use of antithrombotic remains an intractable clinical problem on account of the complicated vascular endothelial condition, chronic atherosclerotic disease, etc[1]. Additionally, effective cardiac rehabilitation can be viewed as another improvement of prognosis, which is a form of comprehensive and long-term exercise composed of risk factor analysis, physical activities, mental support, life-style behavior interventions[2-4]. The World Health Organization deems that exercise-based cardiac rehabilitation can influence patient’s physical, psychological and social condition, benefit their quality of life and control potential complications[5]. Furthermore, safe exercise at different intensity can affect the training endurance, oxygen capacity and intervention effects [6].

It is recommended that moderate-intensity continuous training(MICT) at the intensity of 50% to 75% heart rate[7] or vigorous-intensity interval exercise are both beneficial to maintain people’s health and prevent from the occurrence of disease[8, 9]. In spite of these advantages, around 30% adults fail to meet the demand because of lacking of time and hard to persist[10]. More importantly, the long period and complexity of exercise lead to patients giving up these activities [11]. Therefore, the high-intensity interval training (HIIT) is referred as an alternative cardiac rehabilitation to medium/low intensity activities, which is short-time high oxygen-consumption exercise interphase with periods of low-intensity training or rest for recovery[8]. To be specific, a maximal or symptom-limited exercise test with the highest heart rate ranging from 85% to 100% is the most common HIIT method.

As for the intervention effects of the two approaches, systematic reviews indicate HIIT improved cardiopulmonary function, blood glucose, lipid and cholesterol profiles, and some inflammatory markers in patients with chronic disease[12-14]. And, the severity of anxiety and depression are markedly declined after HIIT[15, 16]. Especially for CAD patients, compared to MICT, HIIT is more predominant on exercise capacity and quality of life. Despite HIIT outperforms MICT in physical and psychological status for CAD patients, some indicators of cardiac performance were not included in the published studies no matter by direct or indirect ways[17]. Besides these uncertainties, some performance variables of aerobic capacity in cardiac patients showed no difference in terms of systolic or diastolic blood pressure, maximal heart rate, peak minute ventilation and left ventricular ejection fraction[18, 19]. Although the effect of HIIT was gradually explicit, little is explored about the role and the validity of HIIT on patients following PCI. And, combined with regular exercise, CAD patients after PCI recover better than PCI alone[3]. The objective of this study is to analyze current research concerning the effect of HIIT in post-PCI patients.

## 2. Methods

This meta-analysis was investigated according to the Preferred Reporting Items for Systematic Review and Meta-analysis(PRISMA)2009 Checklist[20].

### 2.1 Search strategy

Relevant electronic databases were searched such as Cochrane Library, Web of Science Core Collection, Medline, EMbase, PubMed, China National Knowledge Infrastructure (CNKI) and SinoMed from the inception to March 24, 2020. Those databases were retrieved by two researchers using the following strategies independently. Keywords were set as: percutaneous coronary intervention/coronary arteriography/stent implantation/stent placement/drug-eluting stent/percutaneous transluminal coronary angioplasty, and high intensity interval training/high-intensity interval training/high interval training/high interval exercise/HIIT/high intensity interval exercise/intensity aerobic exercise/aerobic exercise. The reference lists of all included papers or related systematic review and meta-analysis were checked to prevent the omissions.

### 2.2 Eligibility Criteria

Studies were included according to the following criteria: (1) patients diagnosed with CAD who underwent PCI; (2) patients in experimental group were mainly treated with HIIT, while patients in control group received usual care or MICT respectively; (3) papers published in English or Chinese; (4) randomized controlled trials (RCTs). Studies were excluded if: (1) literature review, letters, or commentaries, case report, patent, meeting, editorial, and animal trials; (2) the full text could not be accessed in public databases. (3) no efficient data can be extracted; (4) duplicated publications.

### 2.3 Study selection

EndNote X7, the Thomson research software was used screen out the duplicates. Then, titles, topics, abstracts and full text were checked orderly by two researchers (XY Z and DM X) according to the eligible criteria separately. Once any conflicts arose, they sought consultation with a superior researcher.

### 2.4 Data extraction and quality assessment

A form was designed to extracting the basic information. If any data were missing, we would e-mail the corresponding author. The Cochrane handbook for Systematic Reviews of Intervention 5.1.0 was used to appraise the quality of eligible studies in terms of 7 items: randomization, concealment, blind of participation, researcher and data analyst, selective reporting, incomplete outcome data and other biases though judging as “high risk”, “unclear risk” and “low risk”.

### 2.5 Statistical analysis

The authors conducted meta-analysis using Review Manager software (version5.3; Copenhagen, Denmark: the Nordic Cochrane Center, the Cochrane Collaboration, 2103). Considering the continuous outcome, standardized mean difference (SMD) effect size and 95% confidence intervals (CI) were selected for analysis. The heterogeneity of overall effect size was quantified with I^2^ statistic. With and P>0.1, a fixed-effect model was adopted if I^2^<50%,which signify low heterogeneity [21]. Subgroup analysis was used if there exited significant differences between the groups.

## 3. Results

### 3.1 Search results

Based on the eligibility criteria, 314 articles were retrieved though the literature search process. After the elaborative examination and discussion, 6 publications [22-27] were deemed applicable for meta-analysis. The following chart displays the search process and the reason for removing (Figure 1).

**Figure 1.**
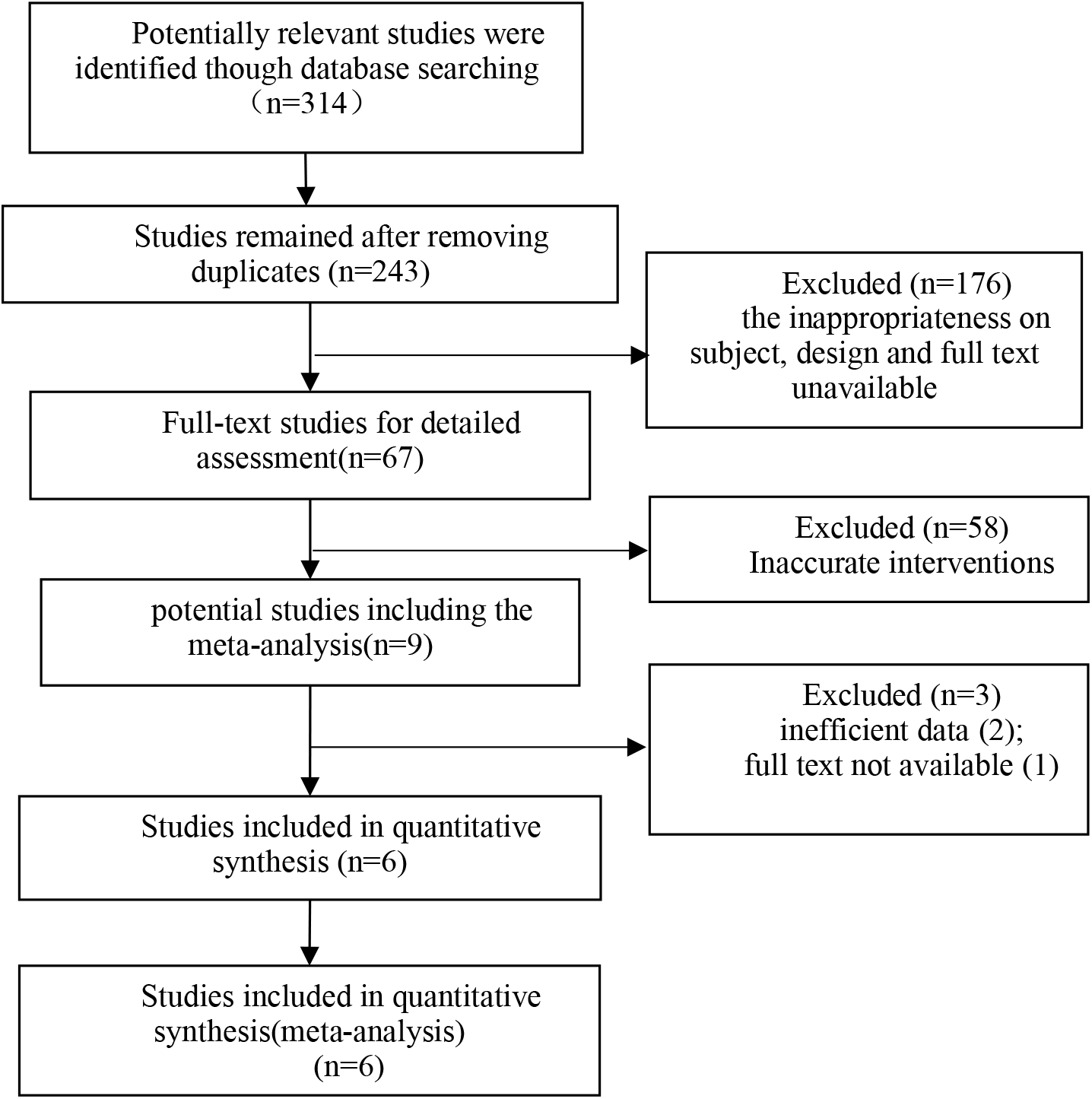
Flow chart of study selection process

### 3.2 Study characteristics

This meta-analysis encompassed 6 RCTs with 247 patients (114 in experimental group and 133 in control group). The baseline characteristics were displayed in the Table 1. Patients after successful PCI were randomized into experimental group conducted with HIIT program and control group underwent less intensive exercise. The common outcomes could consist of three aspects: cardiopulmonary function weighed by left ventricular ejection function (LVEF), peak oxygen take (VO_2peak_) and heart rate (HR), lipid profiles such as high-density lipoprotein (HDL), low-density lipoprotein (LDL) and triglycerides (TGs), and late luminal loss (LLL).

**Table 1:**
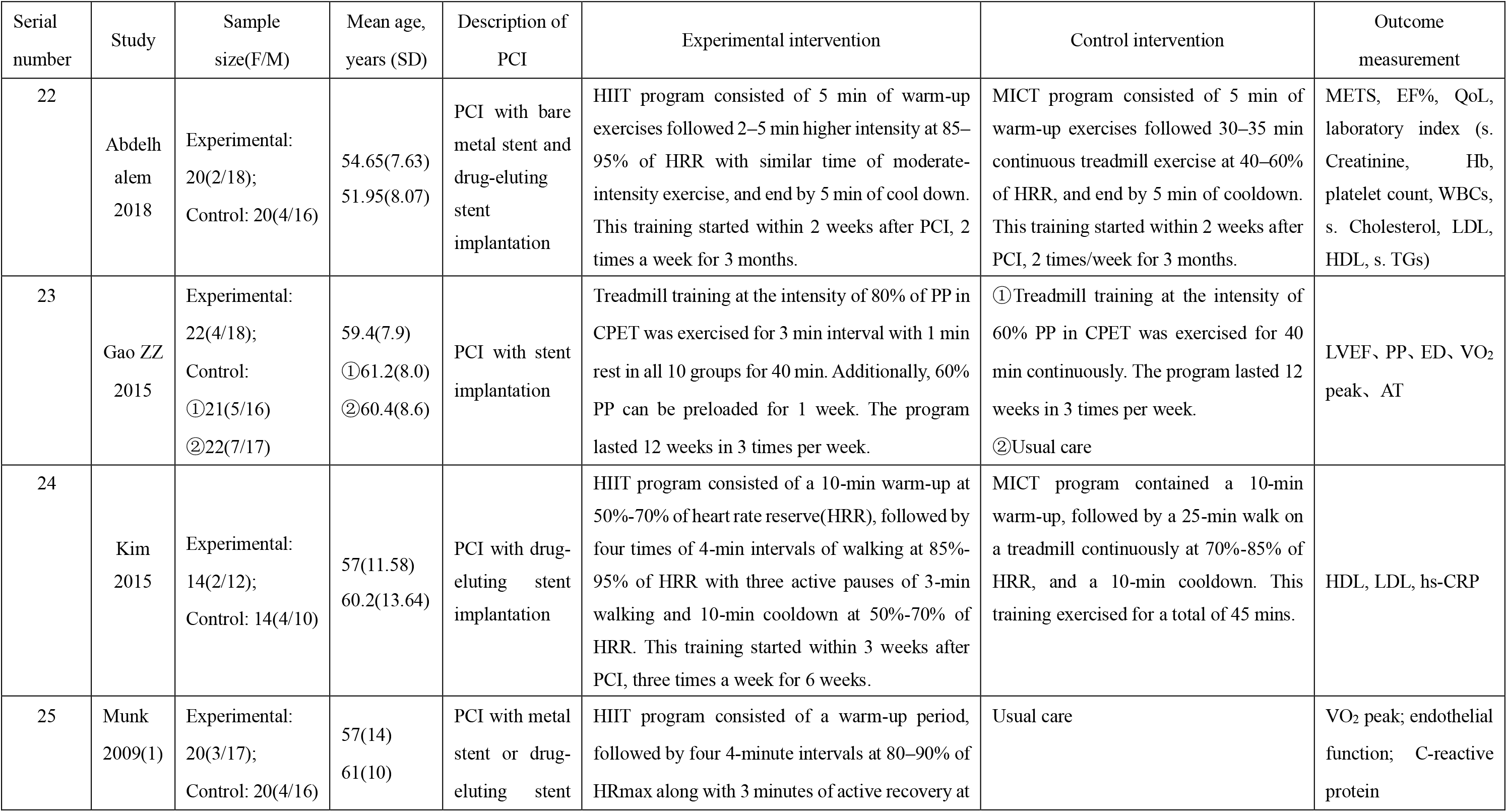
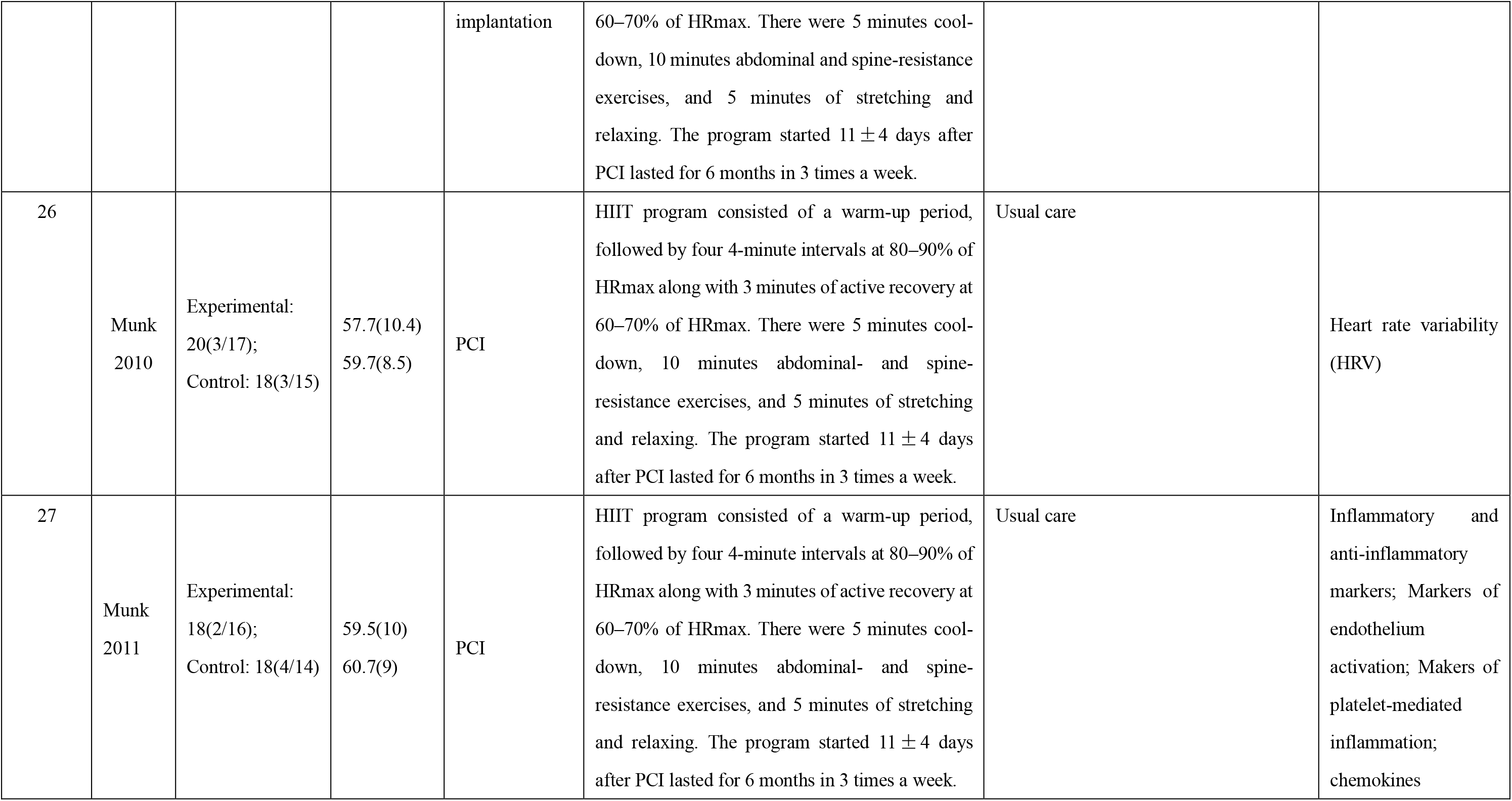
Characteristics of included studies

| Serial number | Study | Sample size(F/M) | Mean age, years (SD) | Description of PCI | Experimental intervention | Control intervention | Outcome measurement |
| --- | --- | --- | --- | --- | --- | --- | --- |
| 22 | Abdelhalem 2018 | Experimental: 20(2/18);<br>Control: 20(4/16) | 54.65(7.63)<br>51.95(8.07) | PCI with bare metal stent and drug-eluting stent implantation | HIIT program consisted of 5 min of warm-up exercises followed 2–5 min higher intensity at 85–95% of HRR with similar time of moderate-intensity exercise, and end by 5 min of cool down. This training started within 2 weeks after PCI, 2 times a week for 3 months. | MICT program consisted of 5 min of warm-up exercises followed 30–35 min continuous treadmill exercise at 40–60% of HRR, and end by 5 min of cooldown. This training started within 2 weeks after PCI, 2 times/week for 3 months. | METS, EF%, QoL, laboratory index (s. Creatinine, Hb, platelet count, WBCs, s. Cholesterol, LDL, HDL, s. TGs) |
| 23 | Gao ZZ 2015 | Experimental: 22(4/18);<br>Control: ①21(5/16)<br>②22(7/17) | 59.4(7.9)<br>①61.2(8.0)<br>②60.4(8.6) | PCI with stent implantation | Treadmill training at the intensity of 80% of PP in CPET was exercised for 3 min interval with 1 min rest in all 10 groups for 40 min. Additionally, 60% PP can be preloaded for 1 week. The program lasted 12 weeks in 3 times per week. | ①Treadmill training at the intensity of 60% PP in CPET was exercised for 40 min continuously. The program lasted 12 weeks in 3 times per week.<br>②Usual care | LVEF, PP, ED, VO<sub>2</sub> peak, AT |
| 24 | Kim 2015 | Experimental: 14(2/12);<br>Control: 14(4/10) | 57(11.58)<br>60.2(13.64) | PCI with drug-eluting stent implantation | HIIT program consisted of a 10-min warm-up at 50%-70% of heart rate reserve(HRR), followed by four times of 4-min intervals of walking at 85%-95% of HRR with three active pauses of 3-min walking and 10-min cooldown at 50%-70% of HRR. This training started within 3 weeks after PCI, three times a week for 6 weeks. | MICT program contained a 10-min warm-up, followed by a 25-min walk on a treadmill continuously at 70%-85% of HRR, and a 10-min cooldown. This training exercised for a total of 45 mins. | HDL, LDL, hs-CRP |
| 25 | Munk 2009(1) | Experimental: 20(3/17);<br>Control: 20(4/16) | 57(14)<br>61(10) | PCI with metal stent or drug-eluting stent | HIIT program consisted of a warm-up period, followed by four 4-minute intervals at 80–90% of HRmax along with 3 minutes of active recovery at | Usual care | VO<sub>2</sub> peak; endothelial function; C-reactive protein |
| | | | | implantation | 60–70% of HRmax. There were 5 minutes cool-down, 10 minutes abdominal and spine-resistance exercises, and 5 minutes of stretching and relaxing. The program started $11 \pm 4$ days after PCI lasted for 6 months in 3 times a week. | | |
| 26 | Munk<br>2010 | Experimental:<br>20(3/17);<br>Control: 18(3/15) | 57.7(10.4)<br>59.7(8.5) | PCI | HIIT program consisted of a warm-up period, followed by four 4-minute intervals at 80–90% of HRmax along with 3 minutes of active recovery at 60–70% of HRmax. There were 5 minutes cool-down, 10 minutes abdominal- and spine-resistance exercises, and 5 minutes of stretching and relaxing. The program started $11 \pm 4$ days after PCI lasted for 6 months in 3 times a week. | Usual care | Heart rate variability (HRV) |
| 27 | Munk<br>2011 | Experimental:<br>18(2/16);<br>Control: 18(4/14) | 59.5(10)<br>60.7(9) | PCI | HIIT program consisted of a warm-up period, followed by four 4-minute intervals at 80–90% of HRmax along with 3 minutes of active recovery at 60–70% of HRmax. There were 5 minutes cool-down, 10 minutes abdominal- and spine-resistance exercises, and 5 minutes of stretching and relaxing. The program started $11 \pm 4$ days after PCI lasted for 6 months in 3 times a week. | Usual care | Inflammatory and anti-inflammatory markers; Markers of endothelium activation; Makers of platelet-mediated inflammation; chemokines |

### 3.3 Quality assessment

The results of quality assessment were listed in detail in figure 2. 2 trials involved the methods of randomization and concealed the way of allocation[23, 25]. But, only 1 trial[25] reported the blinding of participants and personnel. The incomplete, selective report, as well as other bias were appraised in low risk. The amount of “low risk” accounted for 64% in all the checklists.

**Figure 2.**
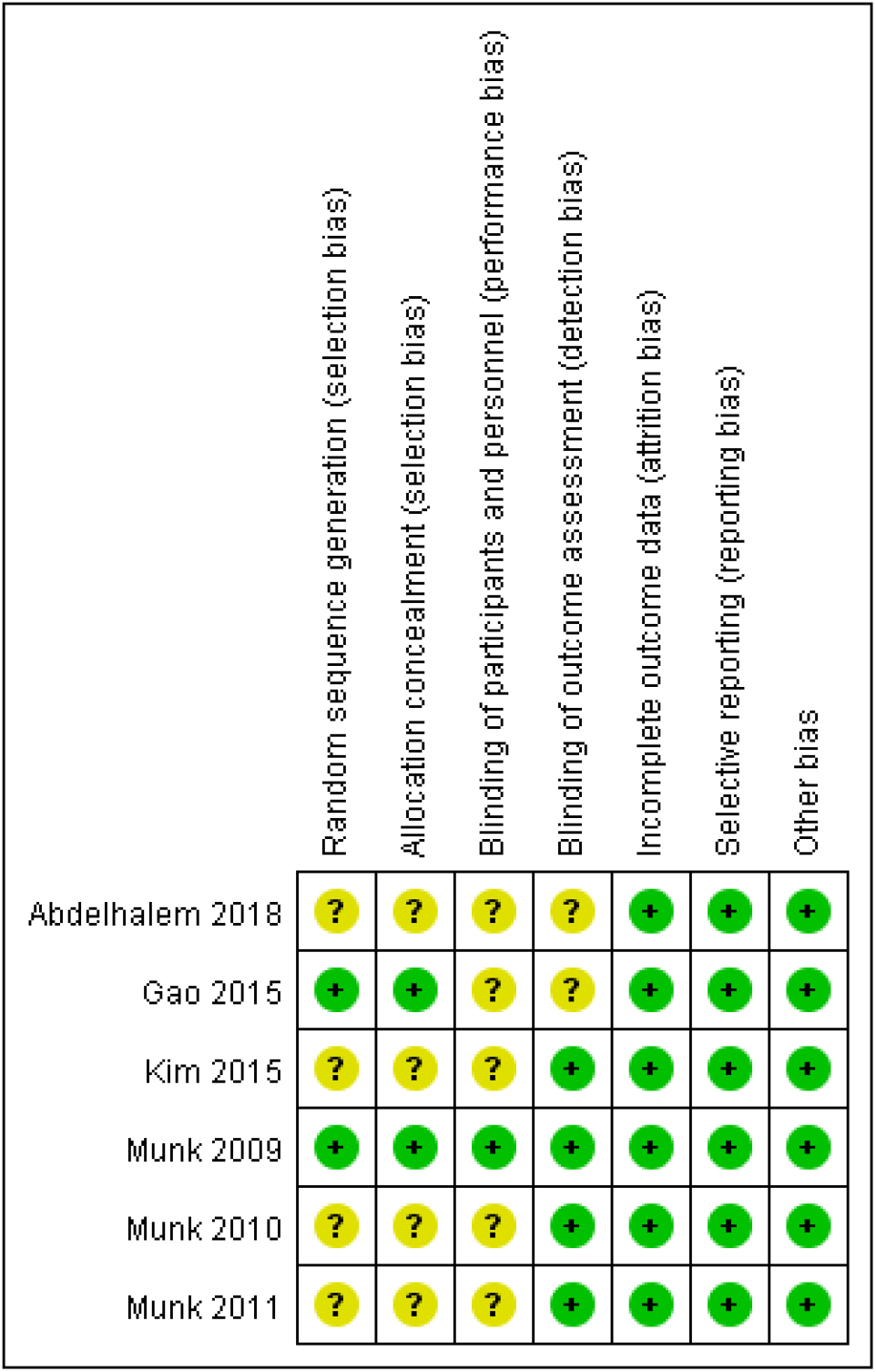
Quality assessment summary for each eligible study

### 3.4 the results of meta-analysis

#### 3.4.1 Left ventricular ejection function (LVEF)

2 RCTs[22, 23] (n=105 patients) reproted the LVEF of post-PCI patients underwent HIIT and less intensive exercise. Considering the participants were randomizedly divided into three groups in the study of Gao[23], the included sample was formed into HIIT vs MICT (Gao 2009(1)) and HIIT vs usual care(Gao 2009(2)). According to the different measurements of control group, the effect of HIIT vs MICT and HIIT vs usual care were estimated in subgroup analysis. The meta-analysis showed the two groups had better homogeneity (I^2^=3%, P=0.36) in the fixed-effect model(Figure 3). HIIT had a statistically significant effect on elevating LVEF (SMD=0.38, 95%CI[0.03, 0.73], p=0.03).

**Figure 3.**
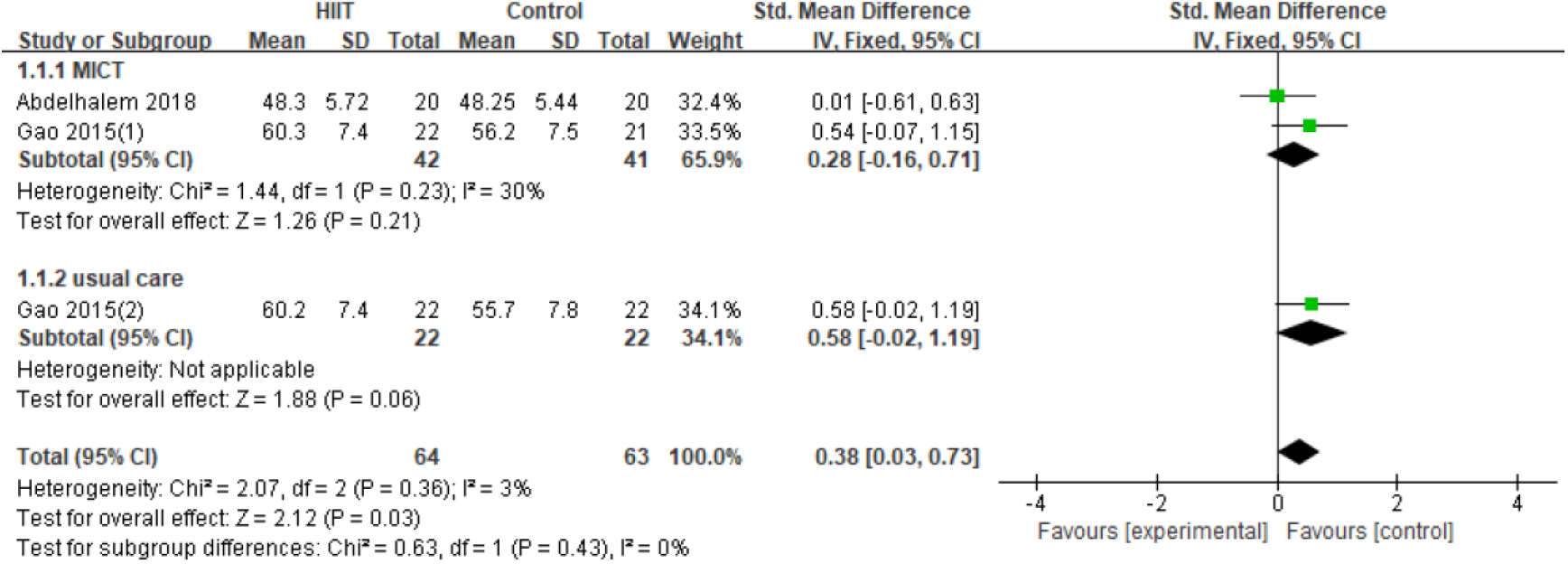
LVEF between two groups

#### 3.4.2 Peak oxygen uptake (VO_2peak_)

3 RCTs[23-25] (n=133 patients) reproted the VO_2peak_ of patients received HIIT and less intensive exercise after PCI. The meta-analysis demonstrated that there did not exist statistical heterogeneity between the two groups (I^2^=0%, P=0.52) in the fixed-effect model(Figure 4). HIIT had a statistically advantegous effect on increasing VO_2peak_ (SMD=0.94, 95%CI[0.61, 1.28], p<0.01).

**Figure 4.**
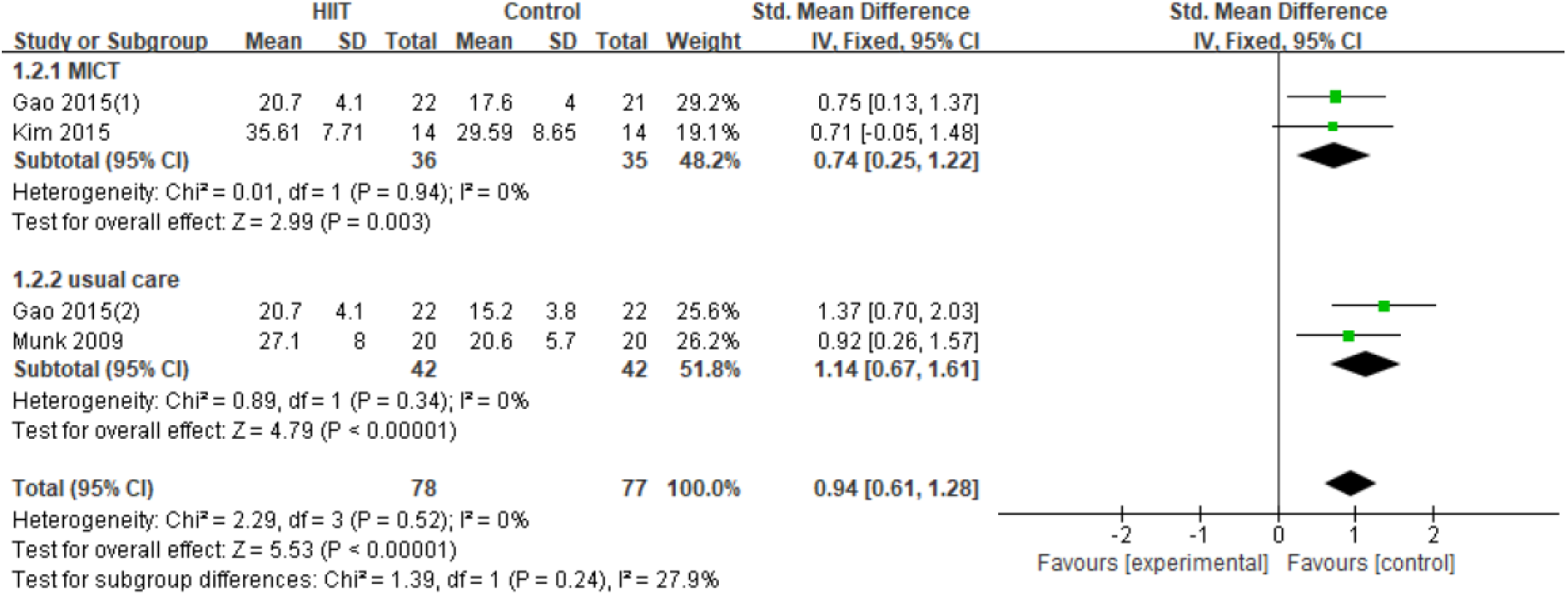
VO_2peak_ between two groups

#### 3.4.3 Heart rate (HR)

3 RCTs[24-26] (n=106 patients) explored patients’ HR in different times. There was no statistical heterogeneity between the experimental and control groups (I^2^=0%, P=0.62) in the fixed-effect model(Figure 5). Compared with regular exercise, HIIT had no significant effect on changing HR (SMD=-0.04, 95%CI[-0.29, 0.21], p=0.73).

**Figure 5.**
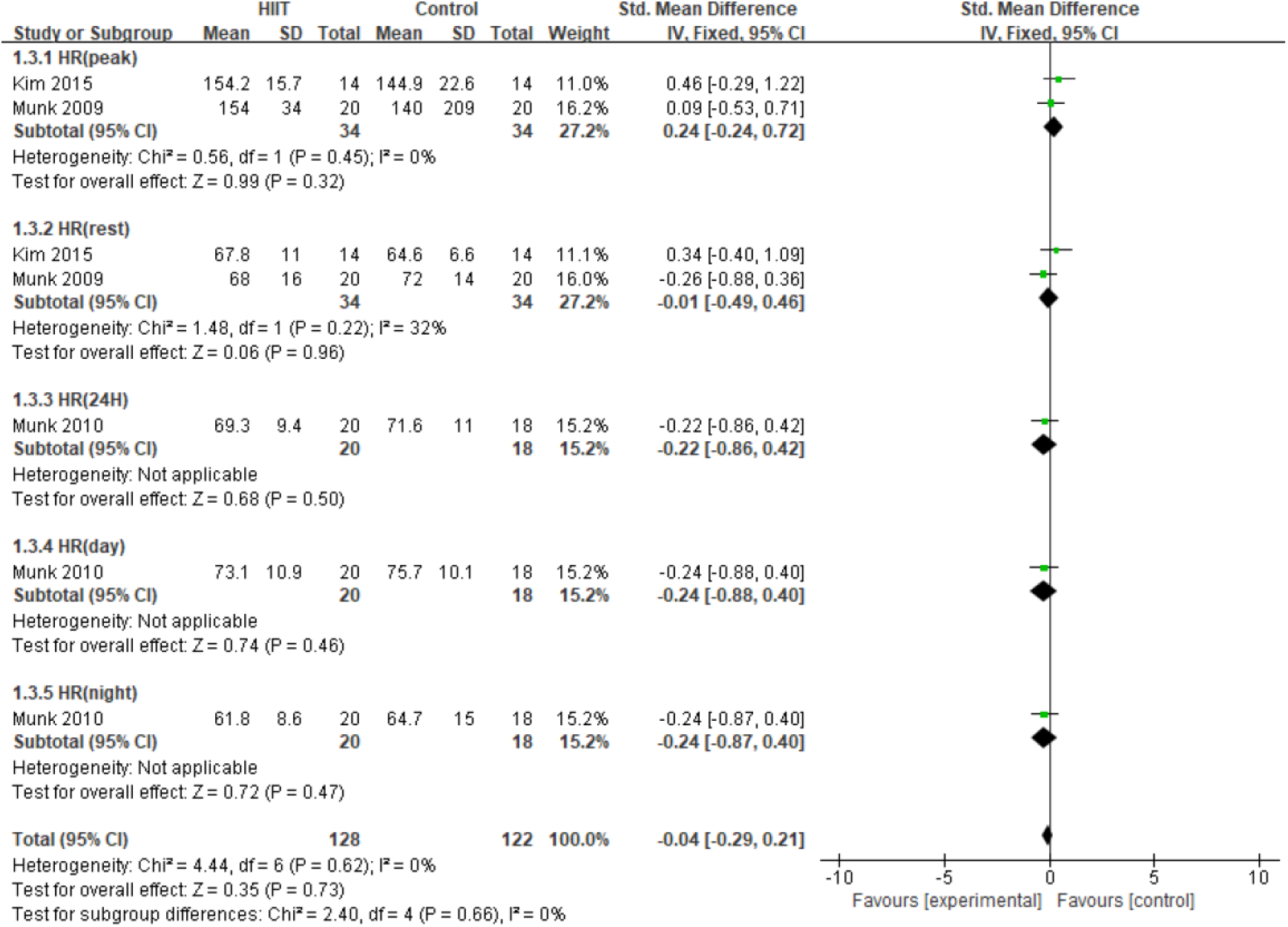
HR between two groups

#### 3.4.4 Lipid profiles

2 RCTs[22, 24] (68 participants) reported the serum level of lipid profiles in the case of two interventions. The result of meta-analysis exhibited no heterogeneity in the level of HDL (I^2^=0%, P=0.87), but a little heterogeneity in LDL and TGs (I^2^=29%, P=0.24) in the fixed-effect model (Figure 6, 7). Hence, HIIT had an weak impact on improving the serum level of HDL (SMD=0.55, 95%CI[0.06, 1.03], p=0.03), and has no effect on LDL and TGs (SMD=-0.05, 95%CI[-0.40, 0.30], p=0.77).

**Figure 6.**
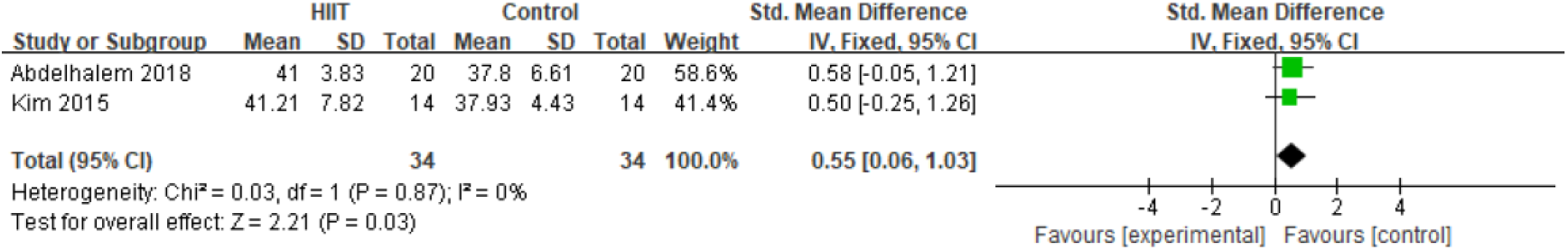
the serum level of HDL between two groups

**Figure 7.**
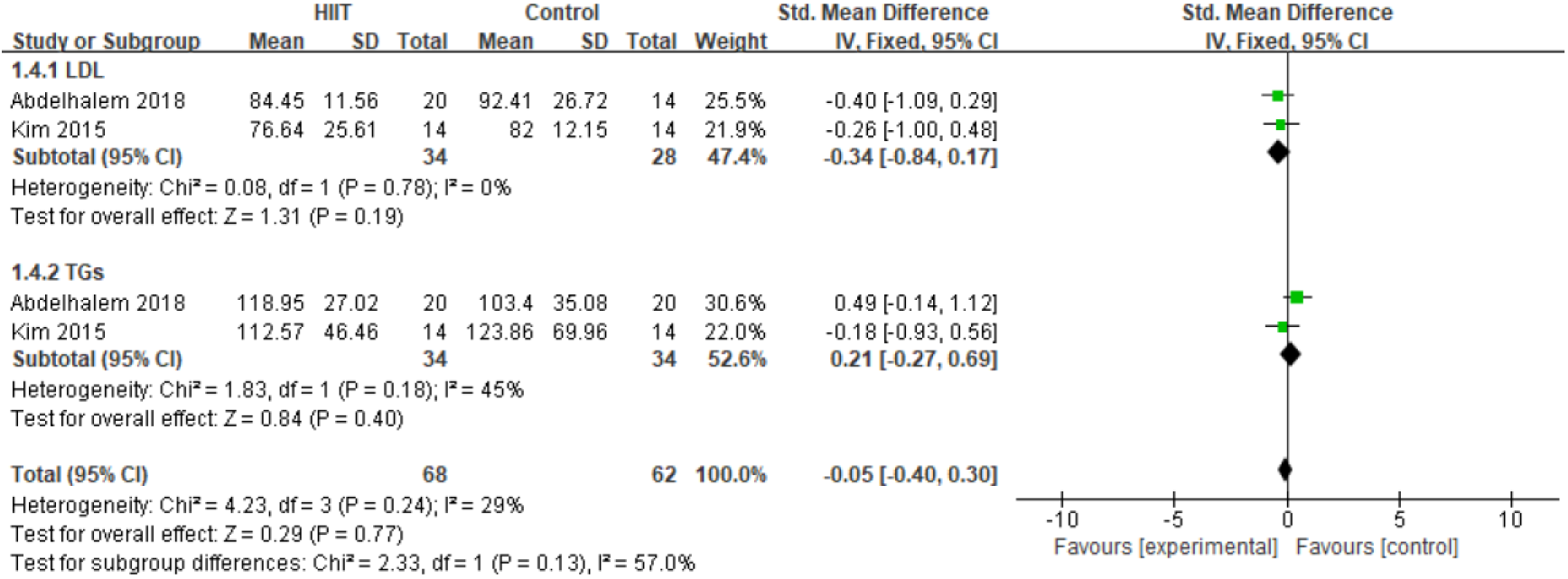
the serum level of LDL and TGs between two groups

#### 3.4.5 late luminal loss (LLL)

Due to the employment of coronary angiography at 6 months after PCI, 2 RCTs[25, 27] (n=76 patients) reproted the LLL of stented coronary artery in patients received different types of exercise. With regard to bare metal stent (BMS) and drug-eluting stent (DES), Munk[25] seperately discussed the LLL of patients implanted with two kinds of stents. The meta-analysis showed no statistical heterogeneity (I^2^=0%, P=0.57) in fixed-effect model(Figure 8). HIIT had a significant effect on shrinking LLL (SMD=-0.65, 95%CI[−1.07, −0.23], p<0.01).

**Figure 8.**
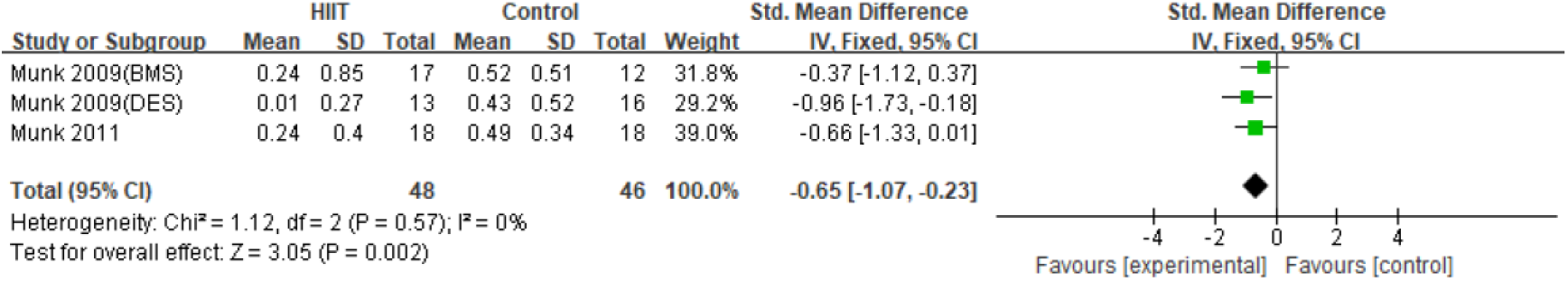
the LLL between two groups

## 4. Discussion

According to the meta-analysis of 6 articles exploring the HIIT program and less intensive exercise in patients diagnosed with CAD undergoing PCI, the results demonstrate that HIIT is an effective intervention to perfect cardiopulmonary function, especially for LVEF and peak oxygen uptake. Besides, late luminal loss is significantly smaller in patients with HIIT program than that with less intensive exercise. However, synthesis of available data also shows that compared with less intensive exercise, HIIT has no significant effect on adjusting HR in patients after PCI whenever during exercise or rest, as well as on the day or night[24-26]. In addition, the serum level of LDL and TGs haven’t obviously changed between two groups. While the serum level of HDL has slightly raised in HIIT group.

### 4.1 Cardiopulmonary function

Compared with no extra exercise in patients, the regular-intensity rehabilitation is beneficial to improve ejection fraction and myocardial contractility in post-PCI patients [3]. In the light of present studies, we deeply investigated that less intensive exercise, even moderate-intensity exercise has not achieved a better effect than HIIT in terms of increasing the levels of LVEF and VO_2peak_ in patients after PCI, which indicates the intensity of exercise program exerts different influence on cardiopulmonary function. Additionally, Munk et al[25] found that ventilatory threshold and maximal workload have greatly increased in post-PCI patients underwent HIIT project. While, the results of this meta-analysis show no delectable changes in peak or resting heart rate between two groups. The functional capacity measured by metabolic equivalents had no statistically significant difference between the HIIT and MICT groups[22]. It has been proved that prescription of HIIT should be given priority to recommend for patients with CAD or high cardiovascular risk who require cardiac rehabilitation in contrast with less intensive exercise[28].

### 4.2 Lipid profiles

When it comes to lipid profiles, this research concludes serum levels of LDL and TGs haven’t markedly improved. Along with HDL cholesterol has raised in HIIT program. This finding has a bit discrepancy with the previous systematic review about healthy people or with physical health complications retrieved by Martland et al[29] that lipid and cholesterol profiles hardly vary with whether the HIIT prescription implemented or not. As the results found in Liou et al[30], no evident differences were surveyed in the levels of HDL and TGs for CAD patients comparing HIIT and MICT programs. Perhaps this is the subtle secret that meta-analysis considers the integrated difference of confidence interval brought by the sample size, which gives more possibility to do high-quality original research to explore this controversy.

### 4.3 Late luminal loss

Promisingly, the meta-analysis shows that late luminal loss in per stented artery for long-time follow-up is greater decreased in HIIT program regardless of the stent types, which indicating the recovery of vessel diameter is satisfied. From an in-depth analysis, the degree of LLL was actively correlated with inflammatory markers such as interleukin (IL)-6[25, 27]. On the other hand, the reduction of LLL indicated the amelioration of endothelial function in accordance with previous study that the increased flow-mediated dilation, as well as the reduction in C-reactive protein were significantly synergistic with LLL following PCI[25]. To some extent, restenosis caused by multiple factors is possibly related to LLL on account of the complicated mechanism of endothelial denudation and released inflammation factors [31, 32]. Therefore, the effect of HIIT program is obviously favorable to decrease the levels of LLL, sequentially influencing the incidence of restenosis of stented arteries.

### 4.4 Effects of intervention characteristics based on previous research

After the PCI with bare metal stent or drug-eluting stent implanted, exercise rehabilitation becomes the fundamental prophylaxis for reducing the occurrence adverse cardiovascular events and improving cardiopulmonary function[33]. The exercise program usually consists of multiform aerobic exercise including treadmill, bicycle ergometer, running and other modes of exercise in different levels of intensity[34]. Generally, HIIT program conducts an interval protocol covering short-time activities at a minimal of 80% of VO_2peak_ or heart rate alternate with relatively less intensive recovery[27, 35-37]. Recent studies have shown that HIIT is superior to MICT in enhancing peak VO_2_ in patients with heart failure and reducing the total fat mass in patients with myocardial infarction[38, 39]. Excepting for the advantages HIIT program brings, the safety should be considered before training. All training sessions in eligible studies were supervised by professional staff even monitored with technical medical devices. Besides, all the rehabilitation program was started from half a month after PCI. On this basis, participants could be more secure in implementing the training to achieve lasting effect of this program, which was proved by a meta-analysis that exercise would positively affect the cardiac function after this program[40].

## 5. Limitations

There are some limitations to be noted in this analysis. Although the eligible trials are randomized and controlled, only few literatures are included in this analysis and the sample size is small leading to the lack of funnel plot. What’s more, it also brings us to another limitation that the enrolled studies are heterogeneous in terms of the interventions in control group. Compared with high-intensity exercise, participants in control group underwent less-intensity exercise covering the moderate-intensity training or usual care which has been amended by the subgroup analysis. Therefore, it is necessary to develop more original studies about the performance of HIIT program in CAD patients following PCI. More importantly, the intensity in experimental and control group should be precise and regulatory.

## 6. Conclusion

This meta-analysis demonstrates that HIIT program might be favorable for CAD patients after PCI by improving cardiopulmonary function, such as LVEF and VO_2peak_, as well as reducing late luminal loss in per stented arteries. Meanwhile, the HIIT has no advantage for adjusting HR. More interestingly, the serum level of HDL has slightly improved while LDL and TGs have no obvious change. Therefore, owing to the limited number of sample size and included studies, the effect of different intensity training, especially for HIIT program remains to be reevaluated meticulously and comprehensively though conducting higher-quality trials in future research.

## Data Availability

Data were collected from published articles.

## Author contributions

*Xinyue Zhang and Dongmei Xu* jointly identified the research problem, information retrieval and data analysis. Besides, this article was written by Xinyue Zhang.

*Guozhen Sun* played a part in determining this topic.

*Zhixin Jiang* helped to polish this article.

*Jinping Tian and Qijun Shan* were both undertake the design of this project and supervise the process of research to guarantee the authenticity of paper.

## Acknowlegements

We sincerely appreciate all the beneficial comments given by our instructor and members of research group. The authors have no conflicts of interest to declare the content of this literature.

